# Biomarkers’ performance in the SEPSIS-3 era

**DOI:** 10.1101/2023.01.18.23284703

**Authors:** Amanda de la Fuente, Jaime López-Sánchez, Luis Mario Vaquero-Roncero, María Merino García, María Elisa Sánchez Barrado, Miguel Vicente Sánchez-Hernández, Jesús Rico-Feijoo, Luis Muñoz-Bellvís, Rafael González de Castro, Ana P. Tedim, Alicia Ortega, Omar Abdel-lah Fernández, Alejandro Suárez-de-la-Rica, Emilio Maseda, Ignacio Trejo González, Geovanna Liszeth García Carrera, José Miguel Marcos-Vidal, Juan Manuel Nieto Arranz, Carmen Esteban-Velasco, César Aldecoa, Jesús F Bermejo-Martin

## Abstract

**Objective:** the biomarkers’ performance for diagnosis and severity stratification of sepsis has not been properly evaluated anew using the SEPSIS-3 criteria introduced in 2016. We evaluated the accuracy of 21 biomarkers classically tested in sepsis research to identify infection, sepsis, and septic shock in surgical patients classified using SEPSIS-3.

**Methods:** four groups of adult surgical patients were compared: post-surgical patients with no infection, patients with infection but no sepsis, patients with sepsis, and patients with septic shock were recruited prospectively from the surgery departments and surgical ICUs from four Spanish hospital. The area under the curve (AUC) to differentiate between groups was calculated for each biomarker.

**Results:** A total of 187 patients were recruited (50 uninfected post-surgery controls, 50 patients with infection, 47 with sepsis and 40 with septic shock). The AUCs indicated that none of the biomarkers tested was accurate enough to differentiate those patients with infection from the uninfected controls. In contrast, procalcitonin, lipocalin 2, pentraxin 3, IL-15, TNF-α, IL-6, angiopoietin 2, TREM-1, D-dimer and C-reactive protein yielded AUCs > 0.80 to discriminate the patients with sepsis or septic shock from those with no infection. C-reactive protein and IL-6 were the most accurate markers to differentiate plain infection from sepsis (AUC = 0.82). Finally, our results revealed that sepsis and septic shock shared similar profiles of biomarkers.

**Conclusion:** Revaluation in the “SEPSIS-3 era” identified the scenarios where biomarkers do and do not provide useful information to improve the management of surgical patients with infection or sepsis.

## Introduction

Identification of sepsis remains a major challenge to implement prompt treatment in surgical patients suffering from this condition. Correct and quick discrimination between sepsis and surgical related inflammation allows to early implement measures aimed to control the infection source with surgery or antibiotics (1,2). Biomarkers are a potential useful tool to improve sepsis detection, complementary to clinical information and to image and/or microbiological tests, but the information regarding biomarkers must be provided in minutes in order to be useful (3,4). In addition, the emergence of the SEPSIS-3 criteria in 2016 has re-shaped sepsis diagnosis, by proposing a new definition which consider sepsis just those infections causing life-threatening organ failure (5,6). While the introduction of the new SEPSIS-3 criteria has impacted epidemiological studies on sepsis (7–9), how SEPSIS-3 affects the performance of sepsis biomarkers has not been sufficiently studied yet.

In this work, we profiled a large number of biomarkers classically tested in sepsis studies by using a rapid microfluidics-based test, to evaluate their performance regarding identification of infection, sepsis and septic shock in surgical patients.

## Methods

### Study design and patients

Adult patients (≥ 18 years) recruited in the first 24 hours following an abdominal surgery with no infection constituted the uninfected control group. Adult patients with infection, sepsis, or septic shock of abdominal source were recruited prospectively from the surgery departments and surgical intensive care units (ICUs) of the four participating hospitals (Hospital Universitario Río Hortega de Valladolid, Complejo Asistencial Universitario de Salamanca, Complejo Asistencial Universitario de León and Hospital Universitario Marqués de Valdecilla de Santander), between January 2020 and July 2022. Infection was defined according to the US Centers for Disease Control and Prevention National Surveillance Definitions for Specific Types of Infections (10). Sepsis and septic shock were defined using the SEPSIS-3 consensus definitions (5,6). A specific standard survey was employed in the four participating hospitals to collect clinical data along with results of hematological, biochemical, radiological, and microbiological investigations. Healthy controls with similar age and sex characteristics to the patients were recruited from the Centro de Hemoterapia y Hemodonación de Castilla y León (CHEMCYL, Valladolid, Spain).

### Biomarkers profiling

we quantified 20 biomarkers in plasma involved in different biological functions using the Ella-SimplePlex TM system from Biotechnne (San Jose, California, USA) as per manufacturer instructions. The biomarkers studied were the following: Lipocalin-2 (LCN2), Myeloperoxidase (MPO) (Neutrophil degranulation); Intercellular adhesion molecule 1 (ICAM-1), Vascular cell adhesion molecule 1 (VCAM-1), Endothelin-1 (ET-1), Angiopoietin 2 (ANGPT2), Angiopoietin 1 (ANGPT1) (Endothelial dysfunction); D-dimer, Urokinase-type plasminogen activator (uPA) (Coagulation); Interleukin 6 (IL-6), Interleukin 15 (IL-15), Tumoral necrosis factor α (TNF-α), Procalcitonin (PCT), Matrix metalloproteinase 7 (MMP7), Pentraxin 3 (PTX3), TREM-1 (Inflammation); Interleukin 10 (IL-10), Programmed Death-ligand 1 (PD-L1) (immunosuppression / immunomodulation), C-X-C motif chemokine ligand 10 (CXCL10), Interleukin 7 (IL-7) (lymphocyte biology). Serum C-reactive protein (CRP) was measured by particle enhanced immunoturbidimetric assay (e501 Module Analyser, Roche Diagnostics, Meylan, France); limit of detection 0.15 mg/dL.

### Statistical analysis

Statistical analysis was performed using IBM SPSS Statistics 25.0 (SPSS INC, Armonk, NY, U.S.A). The level of significance was set at 0.05. For clinical characteristics of the patients, differences between groups were assessed using the χ2 test for categorical variables. Differences between groups for continuous variables and protein levels were assessed with the Kruskal–Wallis test. The accuracy of protein levels to differentiate between groups of patients was studied by calculating the area under the receiver operating characteristic curve (AUC). The optimal operating point (OOP) was calculated on the curve as previously described (11).

## Results

Our study involved 187 patients, 50 uninfected post-surgery controls, 50 patients with infection without sepsis, 47 with sepsis and 40 with septic shock. Patients with infection were significantly younger than those in the other groups. Proportion of men to women were similar in all the compared groups. Patients with sepsis and septic shock had more frequently hypertension and chronic cardiac disease. Septic shock patients were the most severe as evidenced by their SOFA scores at admission and stayed longer at the hospital. None of the patients of the surgical control group or in the infection group died during hospitalization, compared with 7 out of 47 (14.9 %) patients with sepsis and 10 out of 40 (25 %) patients with septic shock (Table S1, Supplementary material). The Kruskall-Wallis test evidenced that patients with sepsis and septic shock showed higher levels of PCT, LCN2, PTX3, IL-15, TNF-α, IL-6, ANGPT2, TREM-1, D-DIMER, CXCL10, VCAM-1, PD-L1 and MMP7 than healthy controls, surgical controls and patients with infection but no sepsis, being the levels of PCT, LCN2, PTX3 and IL-15 the highest in patients with septic shock (Fig 1; Table S2, Supplementary material).

**Figure 1.**
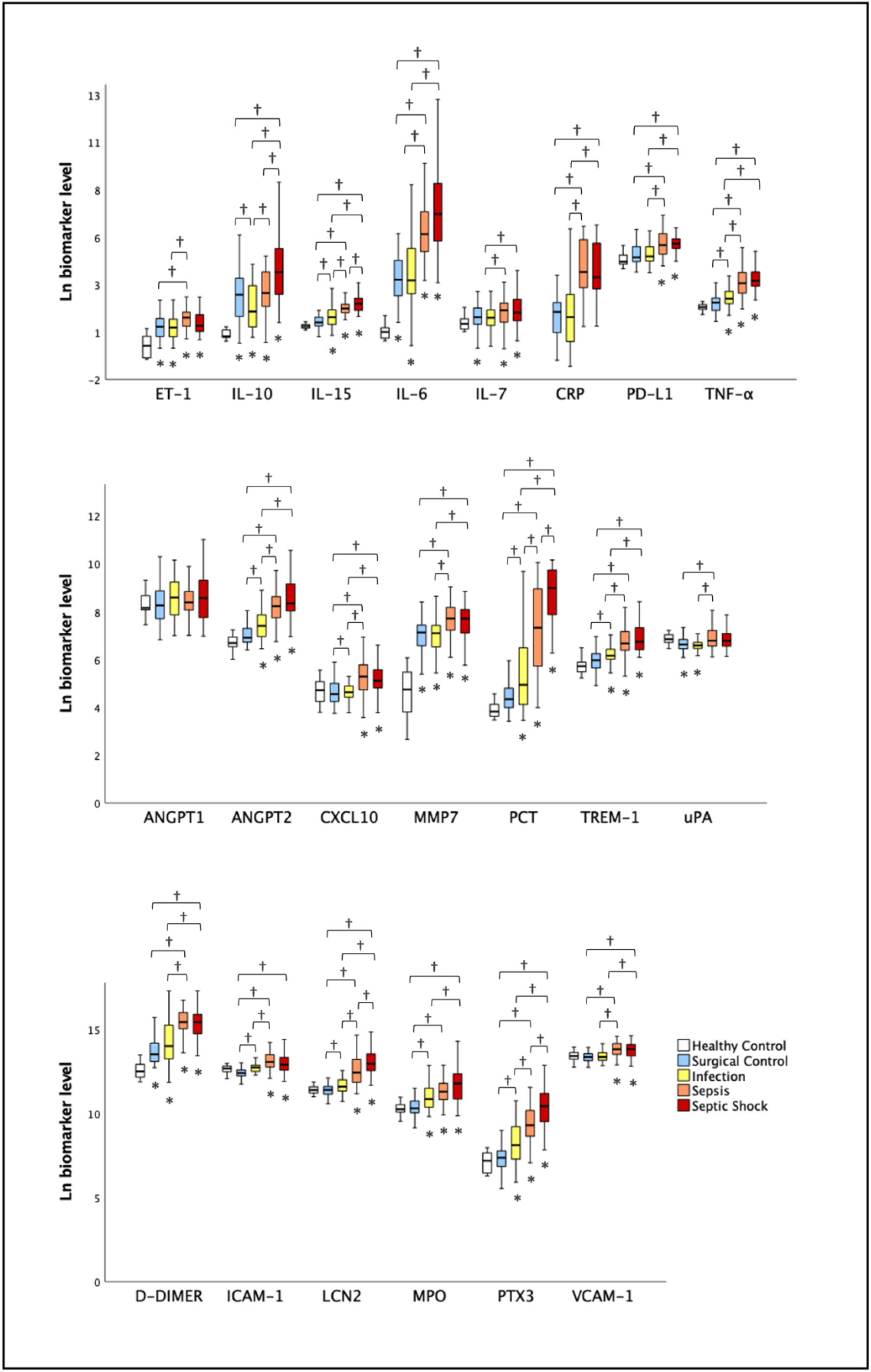
Levels of biomarkers in healthy control, surgical control, infection, sepsis and septic shock groups. Levels are in pg/mL. ET, endothelin; IL, Interleukin; CRP, C reactive protein; PD-L1, programmed death-ligand 1; TNF, tumor necrosis factor; ANGPT, angiopoietin; CXCL, chemokine ligand; MMP, matrix metalloproteinase; PCT, procalcitonin, TREM, triggering receptor expressed on myeloid cells-1; uPA, urokinase-type plasminogen activator; ICAM, intercellular adhesion molecule; VCAM, vascular cell adhesion molecule; PTX, pentraxin; LCN, lipocalin; MPO, myeloperoxidase. \**P* ≤ 0⋅050 *versus* healthy control; † *P* ≤ 0⋅050 (Kruskal – Wallis test).

We next calculated the AUCs for the different biomarkers to discriminate between uninfected post-surgery controls and the patients with infection, sepsis and septic shock (Fig 2; Table S3, Supplementary material). This analysis revealed that none of the biomarkers tested was accurate enough to differentiate patients with a plain infection from post-surgery controls, yielding all AUCs < 0.80. In contrast, PCT, LCN2, PTX3, IL-15, TNF-α, IL-6, ANGPT2, TREM-1, D-DIMER and CRP yielded AUCs > 0.80 to discriminate those patients with sepsis or septic shock from post-surgical patients with no infection (Fig 2; Table S3, Supplementary material). The corresponding OOP are shown Table S4 (Supplementary material). We also evaluated the biomarkers performance to stratify severity. This analysis revealed that CRP and IL-6 were good markers to differentiate plain infection from sepsis, yielding both AUCs of 0.82 for this comparison (Table S3, Supplementary material). The corresponding OOP are shown Table S4 (Supplementary material). In turn, PCT, LCN2, PTX3, IL-15, TNF-α, IL-6, ANGPT2, CRP and IL-10 showed all AUCs > 0.80 to discriminate between infection and septic shock (Table S3, Supplementary material). The corresponding OOP are shown Table S4 (Supplementary material). Finally, our results revealed that sepsis and septic shock shared similar profiles of biomarkers, with none of them yielding AUCs > 0.80 to differentiate between these two conditions (Table S3, Supplementary material).

**Figure 2.**
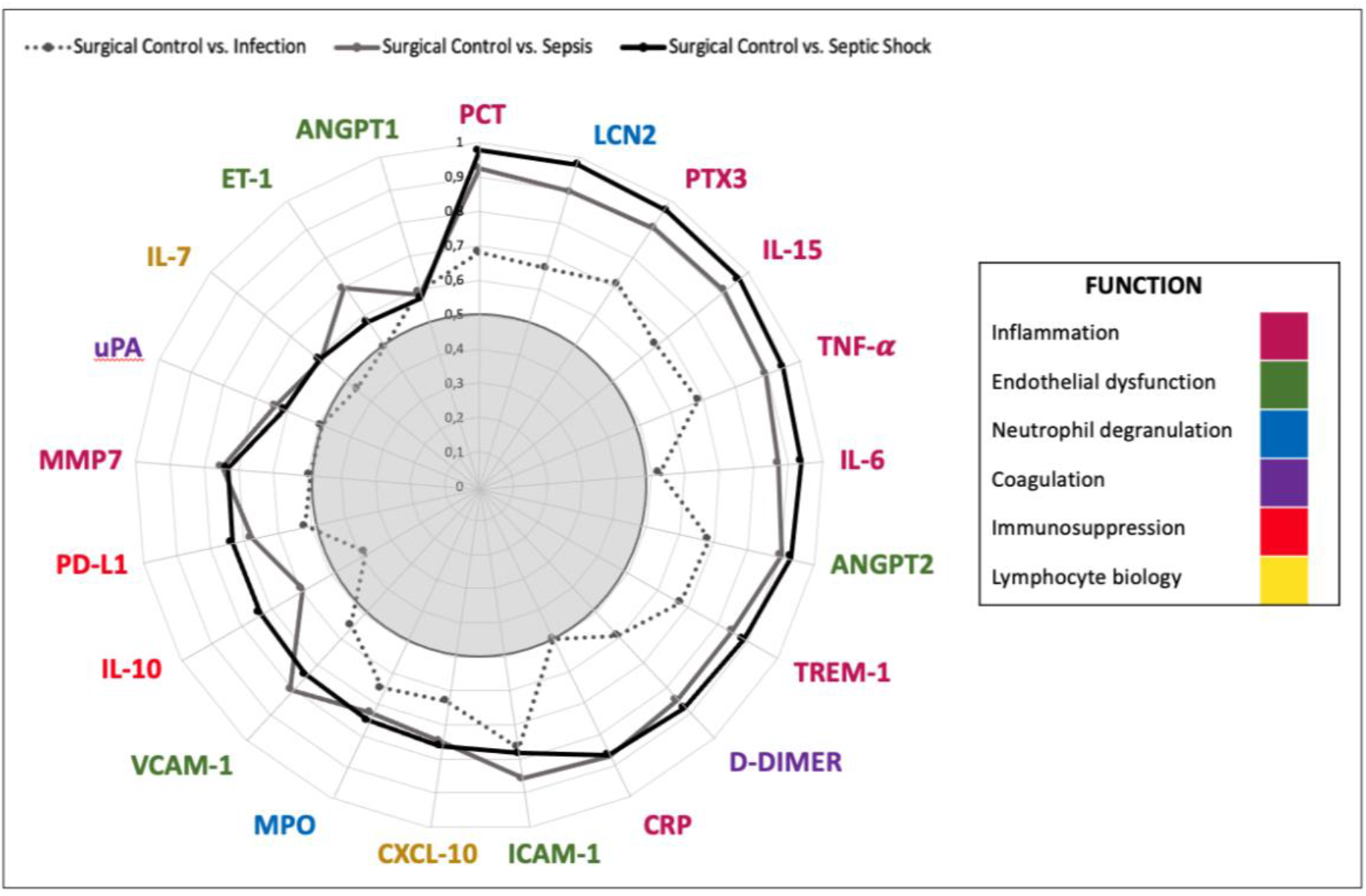
“Big Bang” Plot. AUC to differentiate patients with infection, sepsis, and septic shock from surgical controls. ET, endothelin; IL, Interleukin; CRP, C reactive protein; PD-L1, programmed death-ligand 1; TNF, tumor necrosis factor; ANGPT, angiopoietin; CXCL, chemokine ligand; MMP, matrix metalloproteinase; PCT, procalcitonin, TREM, triggering receptor expressed on myeloid cells-1; uPA, urokinase-type plasminogen activator; ICAM, intercellular adhesion molecule; VCAM, vascular cell adhesion molecule; PTX, pentraxin; LCN, lipocalin; MPO, myeloperoxidase.

## Discussion

Since the introduction of the new SEPSIS-3 criteria in 2016, studies evaluating the performance of biomarkers to diagnose and to stratify sepsis severity are lacking or focused on a limited number of molecules (12–16). Here we evaluated 21 biomarkers involved in different biological functions in sepsis (inflammation, neutrophil degranulation, endothelial dysfunction, coagulation, immunosuppression and lymphocyte biology), and compared their performance to discriminate between surgical patients with no infection, infection with no sepsis, sepsis or septic shock, as defined by SEPSIS-3. Our results evidenced the limitations of the assessed biomarkers to differentiate those patients with infection from those with no infection, revealing that, in absence of significant organ failure, the biological response to an infectious or to a surgical challenge is similar. While these results evidence that the biomarkers tested would not be helpful to better allocate antibiotic treatment in patients with suspected infection when sepsis is absent, we identified in contrast a number of them (PCT, LCN2, PTX3, IL-15, TNF-α, IL-6, ANGPT2, TREM-1, D-DIMER, CRP) which definitively could contribute to quickly identify those patients with sepsis or septic shock and to early implement empiric therapy with wide spectrum antibiotics, along with the other bundles recommended by the surviving sepsis campaign (hemodynamic management, ICU admission, antimicrobial therapy, implemention of any required source control intervention, ventilation and other additional therapies) (17). In turn, CRP and IL-6 were also good candidates to differentiate surgical patients with infection with or without sepsis. This finding is also very important from a translational point of view since quantification of these biomarkers is widely available in hospital settings. Finally, our study revealed that none of the biomarkers evaluated was good enough to differentiate between patients with sepsis and those with septic shock, revealing that both scenarios induce similar alterations in the host response.

A strength of our study is that we employed a next-generation immunoassay based on microfluidics (Ella-SimplePlex) which provides biomarkers levels in less than 90 minutes, which is a reasonable frame time to provide actionable information in patients with suspected sepsis or septic shock. While the limited sample size makes this a pilot study, our results warrant further evaluation of biomarker profiling using Ella-SimplePlex in larger cohorts of patients.

In conclusion, our study re-approached the performance of sepsis biomarkers in the “SEPSIS-3 era”, identifying the scenarios and molecules really adding valuable information to improve the management of surgical patients suffering this deadly condition.

## Data Availability

The datasets generated and/or analysed during the current study are not publicly available since they are still under elaboration for publication by the authors but are available from the corresponding author on reasonable request.

## List of abbreviations

ANGPT2: Angiopoietin 2
AUC: Area under the receiver operating characteristic curve
CXCL10: C-X-C motif chemokine ligand 10
ICAM-1: Intercellular adhesion molecule 1
ICU: Intensive care unit
IL-6: Interleukin 6
IL-7: Interleukin 7
IL-10: Interleukin 10
IL-15: Interleukin 15
MMP7: Matrix metalloproteinase 7
OOP: Optimal operating point
PD-L1: Programmed Death-ligand 1
PCT: Procalcitonin
SOFA: Sepsis related Organ Failure Assessment
TNF-α: Tumor necrosis factor α
TREM-1: Triggering receptor expressed on myeloid cells 1
uPA: Urokinase-type plasminogen activator
VCAM-1: Vascular cell adhesion molecule 1

## Ethics approval and consent to participate

The study was approved by the respective Committees for Ethics in Clinical Research of the three participating hospitals. Methods were carried out in accordance with current Spanish law for Biomedical Research, fulfilling the standards indicated by the Declaration of Helsinki. The study was approved by the Committee for Ethical Research of the coordinating institution, “Comite de Etica de la Investigacion con Medicamentos del Área de Salud de Valladolid Oeste”, code PI142-19. Written informed consent was obtained from patients’ relatives or their legal representative before enrolment.

### Consent for publication

not applicable

### Competing interests

The authors declare that they have no competing interests **Funding:** This study has been funded by Instituto de Salud Carlos III (ISCIII) and co-funded by the European Union: Project “PI19/00590” (JFBM), Sara Borrell program”CD018/0123” (APT) and PFIS program “FI20/00278” (AdF). The funding sources did not play any role in the design of the study and collection, analysis, interpretation of data or writing the manuscript.

### Authors’ contributions

JFBM designed the study. AdlF, JL, LMVR, MMG, MESB, MVSH, JMMV, JRF, LMB, RGdC, APT, AO, ASdR, EM, CEV and CA contributed with patient recruitment and data acquisition. AdlF and AO profiled biomarker levels in plasma. JFBM and AdlF analyzed and interpretated of data and drafted the manuscript. All the authors critically reviewed the article and provided final approval of the version submitted for publication.

## Acknowledgements

The authors thank the nursing teams of the participating clinical services for their continuous support to the research programme. They also thank the Biobanco Hospital Clínico Universitario de Salamanca, for assistance with sample storing, and CHEMCYL (Valladolid, Spain) for providing the samples from healthy controls.

## Notes

### Competing Interest Statement

The authors have declared no competing interest.

### Funding Statement

This study has been funded by Instituto de Salud Carlos III (ISCIII) and co-funded by the European Union: Proyect (PI19/00590), Sara Borrell Research Grant (CD018/0123) (APT), PFIS Research Grant (FI20/00278) (AdF). The funding sources did not play any role in the design of the study and collection, analysis, interpretation of data or writing the manuscript.

### Author Declarations

The study was approved by the Committee for Ethical Research of the coordinating institution, Comite de Etica de la Investigacion con Medicamentos del Area de Salud de Valladolid Oeste, code PI142-19.

