## Supplementary material for "Biomarkers’ performance in the SEPSIS-3 era"

| | **Post-surgery Control (1)** | **Infection (2)** | **Sepsis (3)** | **Septic Shock (4)** | ***p*-value (1 vs. 2)** | ***p*-value (1 vs .3)** | ***p*-value (1 vs. 4)** | ***p*-value (2 vs. 3)** | ***p*-value (2 vs. 4)** | ***p*-value (3 vs .4)** |
|---|---|---|---|---|---|---|---|---|---|---|
| **No. (%)** | | | | | | | | | | |
| | 50 (26.70) | 50 (26.70) | 47 (25.20) | 40 (21.40) | - | - | - | - | - | - |
| **Characteristics** | | | | | | | | | | |
| Age (years) [median (IQR)] | 61.50 (15.00) | 53.00 (37.00) | 71.00 (14.00) | 73.00 (20.00) | **0.040** | **< 0.001** | **0.005** | **< 0.001** | **< 0.001** | n.s. |
| Male [n (%)] | 35 (70.00) | 32 (64.00) | 35 (76.10) | 26 (65.00) | n.s. | n.s. | n.s. | n.s. | n.s. | n.s. |
| **Comorbidities, [ n (%)]** | | | | | | | | | | |
| Allergy | 4 (8.00) | 2 (4.10) | 9 (19.60) | 4 (10.00) | n.s. | n.s. | n.s. | **0.018** | n.s. | n.s. |
| Alcoholism | 0 (0.00) | 0 (0.00) | 3 (6.5) | 0 (0.00) | - | n.s. | - | n.s. | - | n.s. |
| Smoking | 3 (6.00) | 5 (10.40) | 5 (10.90) | 5 (12.5) | n.s. | n.s. | n.s. | n.s. | n.s. | n.s. |
| Chronic Cardiac disease | 4 (8.00) | 6 (12.20) | 16 (34.80) | 14 (35.0) | n.s. | **0.001** | **0.002** | **0.020** | **0.022** | n.s. |
| COPD | 3 (6.00) | 1 (2.00) | 9 (19.60) | 2 (5.0) | n.s. | **0.049** | n.s. | **0.009** | n.s. | **0.044** |
| Asthma | 1 (2.00) | 3 (6.10) | 5 (10.90) | 1 (2.50) | n.s. | n.s. | n.s. | n.s. | n.s. | n.s. |
| Obesity | 4 (8.00) | 7 (14.30) | 5 (10.90) | 5 (12.50) | n.s. | n.s. | n.s. | n.s. | n.s. | n.s. |
| Hypertension | 19 (38.00) | 10 (20.40) | 28 (60.90) | 18 (45.00) | n.s. | **0.031** | n.s. | **< 0.001** | **0.031** | n.s. |
| Dyslipidemia | 16 (32.00) | 10 (20.40) | 19 (41.30) | 11 (27.50) | n.s. | n.s. | n.s. | n.s. | n.s. | n.s. |
| Chronic kidney disease | 2 (4.00) | 1 (2.00) | 7 (15.20) | 2 (5.00) | n.s. | n.s. | n.s. | **0.031** | n.s. | n.s. |
| Chronic liver disease | 1 (2.00) | 0 (0.00) | 0 (0.00) | 3 (3.50) | n.s. | n.s. | n.s. | - | n.s. | n.s. |
| Neurological disease | 2 (4.00) | 5 (10.20) | 3 (6.50) | 7 (17.50) | n.s. | n.s. | **0.037** | n.s. | n.s. | n.s. |
| Immunosuppression | 12 (24.00) | 4 (8.20) | 12 (26.10) | 7 (17.50) | **0.049** | n.s. | n.s. | **0.035** | n.s. | n.s. |
| Active malignant neoplasia | 5 (10.00) | 1 (2.00) | 7 (15.20) | 5 (12.50) | n.s. | n.s. | n.s. | **0.031** | n.s. | n.s. |
| Autoimmune disease | 3 (6.00) | 3 (6.10) | 3 (6.50) | 6 (15.00) | n.s. | n.s. | n.s. | n.s. | n.s. | n.s. |
| Chronic gastrointestinal disease | 9 (18.00) | 3 (6.10) | 4 (8.70) | 2 (5.00) | n.s. | n.s. | n.s. | n.s. | n.s. | n.s. |
| Diabetes Mellitus | 9 (18.00) | 4 (8.20) | 14 (30.40) | 9 (22.50) | n.s. | n.s. | n.s. | **0.011** | n.s. | n.s. |
| **SCORES [median (IQR)]** | | | | | | | | | | |
| SOFA Score | 1.00 (2.00) | 0..00 (1.00) | 3.00 (3.00) | 7.00 (6.00) | n.s. | **< 0.001** | **< 0.001** | **< 0.001** | **< 0.001** | **0.012** |
| **Complications [n (%)]** | | | | | | | | | | |
| Acute respiratory distress | 0 (0.00) | 0 (0.00) | 5 (11.60) | 4 (10.50) | - | **0.013** | **0.019** | **0.015** | **0.021** | n.s. |
| Pulmonary embolism | 0 (0.00) | 0 (0.00) | 0 (0.00) | 1 (2.60) | - | - | n.s. | - | n.s. | n.s. |
| Cardiogenic pulmonary edema | 0 (0.00) | 0 (0.00) | 1 (2.30) | 0 (0.00) | - | n.s. | - | n.s. | - | n.s. |
| Heart arrhythmia | 0 (0.00) | 0 (0.00) | 5 (11.60) | 7 (18.40) | - | **0.013** | **0.002** | **0.015** | **0.002** | n.s. |
| Heart attack | 1 (2.00) | 0 (0.00) | 1 (2.30) | 3 (7.90) | n.s. | n.s. | n.s. | n.s. | **0.048** | n.s. |
| Anastomosis | 0 (0.00) | 2 (4.20) | 1 (2.30) | 2 (5.90) | n.s. | n.s. | n.s. | n.s. | n.s. | n.s. |
| Post operative haemorrhage | 2 (4.00) | 1 (2.10) | 1 (2.30) | 3 (7.90) | n.s. | n.s. | n.s. | n.s. | n.s. | n.s. |
| Gastrointestinal Bleeding | 1 (2.00) | 1 (2.10) | 4 (9.30) | 2 (5.30) | n.s. | n.s. | n.s. | n.s. | n.s. | n.s. |
| Cerebrovascular Accident | 0 (0.00) | 2 (4.20) | 0 (0.00) | 2 (5.40) | n.s. | - | n.s. | n.s. | n.s. | n.s. |
| Paralytic ileus | 1 (2.00) | 7 (14.60) | 4 (9.30) | 9 (24.30) | **0.023** | n.s. | **0.001** | n.s. | n.s. | n.s. |
| Acute liver failure | 0 (0.00) | 0 (0.00) | 3 (7.10) | 1 (2.60) | - | n.s. | n.s. | n.s. | n.s. | n.s. |
| Acute renal failure | 0 (0.00) | 0 (0.00) | 10 (23.30) | 13 (34.20) | - | **< 0.001** | **< 0.001** | **< 0.001** | **< 0.001** | n.s. |
| Secondary Infections | 1 (2.00) | 7 (14.60) | 15 (34.90) | 16 (44.40) | **0.023** | **< 0.001** | **< 0.001** | **0.024** | **0.002** | n.s. |
| Post-operative delirium | 1 (2.00) | 1 (2.00) | 4 (10.50) | 5 (13.50) | n.s. | n.s. | **0.039** | n.s. | **0.047** | n.s. |

| | | | | | | | | | | |
|---|---|---|---|---|---|---|---|---|---|---|
| Coma | 0 (0.00) | 1 (2.10) | 3 (7.50) | 1 (2.80) | n.s. | **0.050** | n.s. | n.s. | n.s. | n.s. |
| Invasive mechanical ventilation | 3 (6.10) | 2 (4.10) | 16 (36.40) | 25 (67.60) | n.s. | **< 0.001** | **< 0.001** | **< 0.001** | **< 0.001** | **0.005** |
| Vasopressor Therapy | 1 (2.00) | 1 (2.00) | 9 (20.90) | 40(100.00) | n.s. | **0.005** | **< 0.001** | **0.005** | **< 0.001** | **< 0.001** |
| Other Complications | 1 (2.0) | 3 (6.10) | 12 (26.70) | 18 (46.20) | n.s. | **< 0.001** | **< 0.001** | **0.007** | **< 0.001** | n.s. |
| **Time course and outcome** | | | | | | | | | | |
| Hospital stay (days) [median (IQR)] | 5.00 (7.00) | 3.50 (7.00) | 10.00 (9.00) | 17.00 (18.00) | n.s. | **< 0.001** | **< 0.001** | **< 0.001** | **< 0.001** | **0.019** |
| Reintervention [n (%)] | 1 (2.00) | 2 (5.00) | 6 (13.30) | 5 (12.50) | n.s. | **0.035** | **0.047** | n.s. | n.s. | n.s. |
| Hospital mortality [n (%)] | 0 (0.00) | 0 (0.00) | 7 (14.90) | 10 (25.00) | - | **0.005** | **< 0.001** | **0.005** | **< 0.001** | n.s. |

**Table S1. Clinical characteristics.** Statistics: Continuous variables are represented as [median (interquartile range)] and categorical variables as absolute count [(n, (%)]. *P-values* were assessed by using the Kruskal–Wallis test for continuous variables, and Chi-squared test for categorical variables. Significant differences ($p < 0.05$) are shown in bold. Abbreviations: p-value, level of significance; COPD, Chronic obstructive pulmonary disease; SOFA, Sequential Organ Failure Assessment.

| BIOMARKER (pg/mL) | Healthy Control (0) | Post-surgery Control (1) | Infection (2) | Sepsis (3) | Septic Shock (4) | *p* value (0 vs 1) | *p* value (0 vs 2) | *p* value (0 vs 3) | *p* value (0 vs 4) | *p* value (1 vs 2) | *p* value (1 vs 3) | *p* value (1 vs 4) | *p* value (2 vs 3) | *p* value (2 vs 4) | *p* value (3 vs 4) |
|---|---|---|---|---|---|---|---|---|---|---|---|---|---|---|---|
| PCT | 46.10 (37.55) | 77.00 (70.47) | 140.50 (621.30) | 1531.00 (7551.00) | 7801.50 (15002.75) | n.s. | **< 0.001** | **< 0.001** | **< 0.001** | **0.013** | **< 0.001** | **< 0.001** | **< 0.001** | **< 0.001** | **0.015** |
| LCN2 | 87937.50 (87937.50) | 88195.00 (40375) | 112509.00 (86832) | 248492.00 (406693) | 420492.00 (472309) | n.s. | n.s. | **< 0.001** | **< 0.001** | **0.032** | **< 0.001** | **< 0.001** | **< 0.001** | **< 0.001** | **0.032** |
| PTX3 | 1317.50 (1513) | 1652.50 (1812) | 3314.00 (8654) | 11006.00 (23220) | 33662.50 (58389) | n.s. | **0.002** | **< 0.001** | **< 0.001** | **0.003** | **< 0.001** | **< 0.001** | **< 0.001** | **< 0.001** | **0.016** |
| IL-15 | 2.24 (0.48) | 2.74 (1.35) | 3.68 (3.32) | 5.66 (2.97) | 7.30 (4.65) | n.s. | **< 0.001** | **< 0.001** | **< 0.001** | **0.004** | **< 0.001** | **< 0.001** | **< 0.001** | **< 0.001** | **0.039** |
| TNF-α | 6.11 (1.70) | 7.73 (4.82) | 9.88 (6.83) | 22.70 (25.80) | 26.25 (30.22) | n.s. | **< 0.001** | **< 0.001** | **< 0.001** | **0.013** | **< 0.001** | **< 0.001** | **< 0.001** | **< 0.001** | n.s. |
| IL-6 | 1.65 (0.89) | 28.20 (66.20) | 25.20 (124.85) | 256.00 (837.00) | 828.00 (4689.00) | **< 0.001** | **< 0.001** | **< 0.001** | **< 0.001** | n.s. | **< 0.001** | **< 0.001** | **< 0.001** | **< 0.001** | n.s. |
| ANGPT2 | 806.00 (363) | 1061.50 (751) | 1649.00 (1617) | 3759.00 (3396) | 4201.00 (6750) | n.s. | **< 0.001** | **< 0.001** | **< 0.001** | **0.011** | **< 0.001** | **< 0.001** | **< 0.001** | **< 0.001** | n.s. |
| TREM-1 | 313.00 (133.50) | 391.50 (233.50) | 488.00 (241.50) | 769.00 (750.00) | 836.50 (975.50) | n.s. | **0.001** | **< 0.001** | **< 0.001** | **0.014** | **< 0.001** | **< 0.001** | **< 0.001** | **< 0.001** | n.s. |
| D-DIMER | 267184.50 (227604) | 740008.50 | 1202340.50 (3729855) | 4164782.00 (6410155) | 4953066.00 (5900275) | **0.001** | **< 0.001** | **< 0.001** | **< 0.001** | n.s. | **< 0.001** | **< 0.001** | **< 0.001** | **< 0.001** | n.s. |
| ICAM-1 | 318073.50 (115048) | 244903.00 (97307) | 345714.50 (150355) | 464609.00 (394522) | 399232.00 (339565) | n.s. | n.s. | **< 0.001** | **0.027** | **< 0.001** | **< 0.001** | **< 0.001** | **0.005** | n.s. | n.s. |
| CXCL10 | 112.50 (91.22) | 95.15 (80.80) | 119.00 (136.62) | 200.00 (257.00) | 171.00 (185.00) | n.s. | n.s. | **0.003** | **0.004** | **0.025** | **< 0.001** | **< 0.001** | **0.032** | **0.032** | n.s. |
| MPO | 28691.00 (15431) | 33989.00 (35267) | 51266.50 (66091) | 80865.00 (99008) | 130081.50 (184640) | n.s. | **0.002** | **< 0.001** | **< 0.001** | **0.044** | **< 0.001** | **< 0.001** | n.s. | **0.003** | n.s. |
| VCAM-1 | 663610.50 (279274) | 631201.50 | 628307.50 (288468) | 996620.00 (698471) | 1000034.00 (688182) | n.s. | n.s. | **< 0.001** | **0.007** | n.s. | **< 0.001** | **< 0.001** | **< 0.001** | **< 0.001** | n.s. |
| IL-10 | 1.29 (0.702) | 11.85 (25.48) | 4.85 (17.05) | 13.30 (40.73) | 39.55 (125.65) | **< 0.001** | **< 0.001** | **< 0.001** | **< 0.001** | **0.038** | n.s. | **< 0.001** | **< 0.001** | **< 0.001** | **0.015** |
| PD-L1 | 71.75 (46.12) | 87.40 (101.67) | 90.20 (100.32) | 162.00 (200.60) | 177.50 (100.50) | n.s. | n.s. | **< 0.001** | **< 0.001** | n.s. | **0.002** | **< 0.001** | **0.003** | **< 0.001** | n.s. |
| MMP7 | 115.00 (204.45) | 1240.50 (1001.75) | 1212.00 (1007.50) | 2224.00 (2203.00) | 2250.50 (2160.25) | **< 0.001** | **< 0.001** | **< 0.001** | **< 0.001** | n.s. | **< 0.001** | **< 0.001** | **< 0.001** | **< 0.001** | n.s. |
| uPA | 942.50 (362) | 760.00 (315) | 738.00 (264) | 891.00 (669) | 842.00 (538) | **0.003** | **0.002** | n.s. | n.s. | n.s. | **0.020** | n.s. | **0.014** | n.s. | n.s. |
| IL-7 | 2.58 (1.44) | 3.59 (3.80) | 3.46 (3.29) | 5.20 (5.04) | 4.59 (7,06) | **0.025** | n.s. | **< 0.001** | **< 0.001** | n.s. | n.s. | n.s. | **0.020** | **0.024** | n.s. |
| ET-1 | 0.79 (0.89) | 2.17 (2.09) | 2.08 (1.98) | 3.52 (2.63) | 2.28 (2.40) | **< 0.001** | **< 0.001** | **< 0.001** | **< 0.001** | n.s. | **< 0.001** | n.s. | **< 0.001** | n.s. | n.s. |
| CRP | | 5.05 (7.17) | 3.63 (378.73) | 39.41 (200.57) | 29.67 (183.10) | | | | | n.s. | **< 0.001** | **< 0.001** | **< 0.001** | **< 0.001** | n.s. |
| ANGPT1 | 3494.50 (2783) | 3875.50 (4976) | 5410.00 (8007) | 4695 (4379) | 5257.50 (9200) | - | - | - | - | - | - | - | - | - | - |

**Table S2. Biomarker levels across groups**. *P-values* were assessed by using the Kruskal–Wallis test. Significant differences ($p < 0.05$) are shown in bold. ET, endothelin; IL, Interleukin; CRP, C reactive protein; PD-L1, programmed death-ligand 1; TNF, tumor necrosis factor; ANGPT, angiopoietin; CXCL, chemokine ligand; MMP, matrix metalloproteinase; PCT, procalcitonin, TREM, triggering receptor expressed on myeloid cells-1; uPA, urokinase-type plasminogen activator; ICAM, intercellular adhesion molecule; VCAM, vascular cell adhesion molecule; PTX, pentraxin; LCN, lipocalin; MPO, myeloperoxidase.

| BIOMARKER | Post-surgery Control vs. Infection | | | Post-surgery Control vs. Sepsis | | | Post-surgery Control vs. Septic Shock | | | Infection vs. Sepsis | | | Infection vs. Septic Shock | | | Sepsis vs. Septic Shock | | |
|---|---|---|---|---|---|---|---|---|---|---|---|---|---|---|---|---|---|---|
| | **AUC** | **IC [95 %]** | ***p*** | **AUC** | **IC [95 %]** | ***p*** | **AUC** | **IC [95 %]** | ***p*** | **AUC** | **IC [95 %]** | ***p*** | **AUC** | **IC [95 %]** | ***p*** | **AUC** | **IC [95 %]** | ***p*** |
| PCT | 0.68 | [0.57-0.79] | **0.002** | 0.92 | [0.87-0.98] | **< 0.001** | 0.98 | [0.94-1.00] | **< 0.001** | 0.78 | [0.69-0.87] | **< 0.001** | 0.92 | [0.87-0.98] | **< 0.001** | 0.71 | [0.61-0.82] | **0.001** |
| LCN2 | 0.66 | [0.56-0.77] | **0.005** | 0.90 | [0.83-0.96] | **< 0.001** | 0.98 | [0.85-1.00] | **< 0.001** | 0.79 | [0.71-0.88] | **< 0.001** | 0.93 | [0.87-0.98] | **< 0.001** | 0.67 | [0.55-0.78] | **0.008** |
| PTX3 | 0.71 | [0.61-0.81] | **< 0.001** | 0.91 | [0.84-0.97] | **< 0.001** | 0.97 | [0.93-1.00] | **< 0.001** | 0.74 | [0.65-0.84] | **< 0.001** | 0.88 | [0.82-0.95] | **< 0.001** | 0.71 | [0.60-0.82] | **0.001** |
| IL-15 | 0.66 | [0.55-0.77] | **0.006** | 0.91 | [0.85-0.97] | **< 0.001** | 0.97 | [0.92-1.00] | **< 0.001** | 0.72 | [0.62-0.83] | **< 0.001** | 0.82 | [0.73-0.91] | **< 0.001** | 0.69 | [0.58-0.80] | **0.003** |
| TNF-α | 0.68 | [0.58-0.79] | **0.002** | 0.89 | [0.83-0.96] | **< 0.001** | 0.95 | [0.90-1.00] | **< 0.001** | 0.78 | [0.69-0.87] | **< 0.001** | 0.87 | [0.79-0.95] | **< 0.001** | 0.59 | [0.47-0.71] | n.s. |
| IL-6 | 0.53 | [0.41-0.64] | n.s. | 0.87 | [0.80-0.94] | **< 0.001** | 0.94 | [0.89-0.99] | **< 0.001** | 0.82 | [0.73-0.90] | **< 0.001** | 0.89 | [0.83-0.96] | **< 0.001** | 0.66 | [0.54-0.77] | **0.012** |
| ANGPT2 | 0.68 | [0.58-0.79] | **0.001** | 0.90 | [0.84-0.96] | **< 0.001** | 0.93 | [0.88-0.98] | **< 0.001** | 0.79 | [0.70-0.88] | **< 0.001** | 0.84 | [0.76-0.92] | **< 0.001** | 0.60 | [0.48-0.73] | n.s. |
| TREM-1 | 0.68 | [0.57-0.78] | **0.002** | 0.85 | [0.77-0.93] | **< 0.001** | 0.89 | [0.82-0.96] | **< 0.001** | 0.74 | [0.64-0.84] | **< 0.001** | 0.79 | [0.70-0.89] | **< 0.001** | 0.55 | [0.42-0.67] | n.s. |
| D-DIMER | 0.59 | [0.48-0.71] | n.s. | 0.85 | [0.76-0.93] | **< 0.001** | 0.88 | [0.81-0.96] | **< 0.001** | 0.72 | [0.62-0.83] | **< 0.001** | 0.76 | [0.66-0.86] | **< 0.001** | 0.53 | [0.40-0.65] | n.s. |
| CRP | 0.49 | [0.37-0.62] | n.s. | 0.87 | [0.79-0.95] | **< 0.001** | 0.87 | [0.78-0.95] | **< 0.001** | 0.82 | [0.73-0.91] | **< 0.001** | 0.82 | [0.73-0.91] | **< 0.001** | 0.48 | [0.36-0.61] | n.s. |
| ICAM-1 | 0.77 | [0.67-0.86] | **< 0.001** | 0.86 | [0.78-0.93] | **< 0.001** | 0.78 | [0.68-0.88] | **< 0.001** | 0.70 | [0.59-0.80] | **0.001** | 0.60 | [0.48-0.73] | n.s. | 0.43 | [0.31-0.55] | n.s. |
| CXCL10 | 0.63 | [0.52-0.74] | **0.023** | 0.75 | [0.65-0.85] | **< 0.001** | 0.76 | [0.67-0.86] | **< 0.001** | 0.62 | [0.51-0.73] | **0.041** | 0.64 | [0.52-0.76] | **0.023** | 0.49 | [0.37-0.61] | n.s. |
| MPO | 0.65 | [0.54-0.76] | **0.010** | 0.73 | [0.62-0.83] | **< 0.001** | 0.75 | [0.65-0.86] | **< 0.001** | 0.63 | [0.51-0.74] | **0.033** | 0.70 | [0.58-0.82] | **0.001** | 0.61 | [0.49-0.74] | n.s. |
| VCAM-1 | 0.55 | [0.43-0.66] | n.s. | 0.80 | [0.72-0.89] | **< 0.001** | 0.74 | [0.63-0.85] | **< 0.001** | 0.77 | [0.68-0.87] | **< 0.001** | 0.70 | [0.58-0.82] | **0.001** | 0.46 | [0.33-0.58] | n.s. |
| IL-10 | 0.38 | [0.27-0.49] | **0.038** | 0.59 | [0.48-0.70] | n.s. | 0.73 | [0.63-0.84] | **< 0.001** | 0.72 | [0.62-0.82] | **< 0.001** | 0.84 | [0.76-0.92] | **< 0.001** | 0.68 | [0.56-0.79] | **0.005** |
| PD-L1 | 0.52 | [0.40-0.63] | n.s. | 0.68 | [0.57-0.78] | **0.003** | 0.73 | [0.63-0.84] | **< 0.001** | 0.67 | [0.57-0.78] | **0.003** | 0.73 | [0.63-0.84] | **< 0.001** | 0.53 | [0.41-0.65] | n.s. |
| MMP7 | 0.49 | [0.38-0.60] | n.s. | 0.75 | [0.65-0.85] | **< 0.001** | 0.73 | [0.62-0.84] | **< 0.001** | 0.75 | [0.65-0.85] | **< 0.001** | 0.73 | [0.63-0.84] | **< 0.001** | 0.49 | [0.37-0.62] | n.s. |
| uPA | 0.49 | [0.38-0.60] | n.s. | 0.64 | [0.52-0.75] | **0.020** | 0.61 | [0.60-0.83] | n.s. | 0.64 | [0.53-0.76] | **< 0.001** | 0.61 | [0.49-0.73] | n.s. | 0.47 | [0.35-0.59] | n.s. |
| IL-7 | 0.45 | [0.34-0.57] | n.s. | 0.59 | [0.47-0.70] | n.s. | 0.59 | [0.49-0.72] | n.s. | 0.64 | [0.53-0.75] | **0.015** | 0.63 | [0.52-0.75] | **0.030** | 0.51 | [0.38-0.63] | n.s. |
| ET-1 | 0.49 | [0.37-0.60] | n.s. | 0.70 | [0.59-0.80] | **0.001** | 0.58 | [0.47-0.71] | n.s. | 0.70 | [0.59-0.80] | **0.001** | 0.60 | [0.48-0.72] | n.s. | 0.38 | [0.26-0.50] | **0.055** |
| ANGPT1 | 0.59 | [0.48-0.70] | n.s. | 0.58 | [0.47-0.70] | n.s. | 0.57 | [0.46-0.70] | n.s. | 0.48 | [0.36-0.59] | n.s. | 0.49 | [0.37-0.61] | n.s. | 0.51 | [0.38-0.63] | n.s. |

**Table S3**. **AUC to differentiate between patients with infection, sepsis, septic shock, and surgical controls.** Significant differences ($p < 0.05$) are shown in bold. ET, endothelin; IL, Interleukin; CRP, C reactive protein; PD-L1, programmed death-ligand 1; TNF, tumor necrosis factor; ANGPT, angiopoietin; CXCL, chemokine ligand; MMP, matrix metalloproteinase; PCT, procalcitonin, TREM, triggering receptor expressed on myeloid cells-1; uPA, urokinase-type plasminogen activator; ICAM, intercellular adhesion molecule; VCAM, vascular cell adhesion molecule; PTX, pentraxin; LCN, lipocalin; MPO, myeloperoxidase.

| BIOMARKER | Post-surgery Control vs. Sepsis | | | Post-surgery Control vs. Septic Shock | | | Infection vs. Sepsis | | | Infection vs. Septic Shock | | |
|---|---|---|---|---|---|---|---|---|---|---|---|---|
| | **OOP** | **Se (%)** | **Sp (%)** | **OOP** | **Se (%)** | **Sp (%)** | **OOP** | **Se (%)** | **Sp (%)** | **OOP** | **Se (%)** | **Sp (%)** |
| PCT | 180 | 87.2 | 60.0 | 455 | 97.5 | 98.0 | | | | 996 | 90.0 | 88.0 |
| LCN2 | 117493 | 82.6 | 84.0 | 180282.50 | 90.0 | 98.0 | | | | 249145.5 | 85.0 | 92.0 |
| PTX3 | 3916.5 | 85.1 | 84.0 | 5934 | 95.0 | 90.0 | | | | 11029 | 82.5 | 80.0 |
| IL-15 | 3.87 | 83.0 | 88.0 | 4.30 | 95.0 | 96.0 | | | | 6.41 | 72.5 | 86.0 |
| TNF-α | 10.7 | 83.0 | 80.0 | 12.75 | 92.5 | 88.0 | | | | 17.75 | 82.5 | 86.0 |
| IL-6 | 100.95 | 76.6 | 84.0 | 131.5 | 92.3 | 90.0 | 65.6 | 85.1 | 68.0 | 147.5 | 84.6 | 78.0 |
| ANGPT2 | 1948 | 83.0 | 92.0 | 2000.5 | 85.0 | 92.0 | | | | 2896.5 | 77.5 | 78.0 |
| TREM-1 | 526 | 83.0 | 82.0 | 534 | 82.5 | 82.0 | | | | | | |
| D-DIMER | 1804955 | 85.1 | 84.0 | 1832863 | 87.2 | 84.0 | | | | | | |
| CRP | 11.11 | 81.8 | 84.8 | 12.1 | 80.0 | 87.9 | 13.15 | 79.5 | 77.6 | 14.39 | 77.5 | 79.6 |
| IL-10 | | | | | | | | | | 22.8 | 67.5 | 84.0 |

**Table S4. Optimal operating point (OOP) of the biomarkers yielded AUCs $\geq$ 0.80 for each comparison.** Results for biomarker levels are provided as pg/mL. Se, sensitivity; Sp, specificity; PCT, procalcitonin; LCN, lipocalin; PTX, pentraxin; IL, Interleukin; TNF, tumor necrosis factor; ANGPT, angiopoietin; TREM, triggering receptor expressed on myeloid cells; CRP, C reactive protein.
